# Effect of the third dose of BNT162b2 vaccine in quantitative SARS-CoV-2 spike 1-2 IgG antibody titers in healthcare workers

**DOI:** 10.1101/2021.10.20.21265269

**Authors:** Maria Elena Romero-Ibarguengoitia, Diego Rivera-Salinas, Yodira Guadalupe Hernández-Ruíz, Ana Gabriela Armendariz-Vázquez, Arnulfo González-Cantú, Irene Antonieta Barco-Flores, Rosalinda González-Facio, Laura Patricia Montelongo-Cruz, Gerardo Francisco Del Rio-Parra, Mauricio René Garza-Herrera, Jessica Andrea Leal-Meléndez, Miguel Ángel Sanz-Sánchez

## Abstract

**Background:** Vaccination is our main strategy to control SARS-CoV-2 infection. Given a decrease in the quantitative SARS-CoV-2 spike 1-2 IgG antibody titers three months following the second BNT162b2 dose, healthcare workers got a third booster dose after six months of completing the original scheme. This study aimed to analyze quantitative SARS-CoV-2 spike 1-2 IgG antibody titers and safety of the third dose.

**Material and methods:** A prospective longitudinal cohort study included healthcare workers who received a third booster dose after six months of the complete BNT162b2 regimen. We assessed the quantitative SARS-CoV-2 spike 1-2 IgG antibody titers 21-28 days after the first and second dose, three months after the complete scheme, 1-7 days following the third dose, and 21-28 days after the boost.

**Results:** The cohort comprised 168 non-immunocompromised participants of 41(10) years old, 67% being women. The third dose was associated with increasing the quantitative antibody titers, regardless of previous SARS-CoV-2 history. In negative SARS-CoV-2 history, the median (IQR) antibody titers increased from 379 (645.4) to 2960 (2010), while in positive SARS-CoV-2 history, from 590 (1262) to 3090 (2080). The third dose had less number of total side effects compared to the other two shots. The most common side effect after the third BNT162b2 shot was pain at the injection site (n=82, 84.5%), followed by tiredness (n=45, 46.4%), with a mild severity (n=36, 37.1%). Tiredness, myalgias, arthralgias, fever, and adenopathy were proportionally higher following the third dose than the two-dose regimen (p<0.05).

**Conclusion:** The third dose applied after six months of the original BNT162b2 regimen provided a good humoral immune response by elevating the quantitative SARS-CoV-2 spike 1-2 IgG antibody titers. The booster dose was well tolerated with no severe side effects after the additional BNT162b2 dose.

## Introduction

In the first trimester of 2020, The World Health Organization (WHO) recognized the spread of SARS-CoV-2 as a pandemic (1). Its negative and severe impact on society, economy, and health due to its significant morbidity and mortality prioritized vaccine development to control the disease worldwide. Many vaccines were developed for emergency use. The vaccines could develop spike-specific IgG antibodies against SARS-CoV-2 since it has been suggested for its high virus neutralization efficacy (2).

The vaccine type depends on its mechanisms of action. So far, the different SARS-CoV-2 vaccines designs are: mRNA (BioNTech-Pfizer/BNT162b2, Moderna/mRNA-1273), adenovirus viral vector (Oxford-AstraZeneca/ChAdOx1, Gam-COVID-VAC/Sputnik V, Ad26.COV2.S/Jannsen, CanSinoBio/Ad5-nCoV), protein subunit (Novavax/NVX-CoV2373, Medicago CoVLP), whole-cell inactivated virus vaccines (Inova/CoronaVac, Sinopharm/ BBIBP-CorV), and DNA vaccines (INO-4800 and ZyCoV-D) (3).

Pfizer and BioNTech’s vaccine, hereafter referred to as BNT126b2, was the first SARS-CoV-2 vaccine to reveal promising efficacy data. On November 18, 2020, it was revealed to be 95% effective against symptomatic and severe disease (4). As a result, in December 2020, the WHO and the Food and Drug Administration (FDA) authorized this vaccine for emergency use, representing the first SARS-CoV-2 vaccine to receive emergency validation (5,6).

The Centers for Disease Control and Prevention (CDC) and the U.S. Food and Drug Administration (FDA) have authorized a third BNT162b2 vaccine to the following individuals: 1) > 50-year-old people with a medical condition; 2) > 18 years old long-term residents of a healthcare facility; 3) 18-49 years old people with a medical condition; and 4) employees and residents in healthcare facilities at high risk for SARS-CoV-2 exposure and transmission (7,8).

It has been proved that the immunity against SARS-CoV-2 induced through BNT126b2 vaccination had a significant degree of protection. However, the duration of its protective immunity is still unknown. Many ongoing studies focused on public concerns about the safety and efficacy of BNT126b2 over time (9). These studies had reported a significant antibody decrease three and six months post-vaccination on individuals who completed the two-dose regimen of the vaccine (9–11). Besides, new strains of the SARS-CoV-2 virus could be developed if it continues to replicate and transmit, which may become resistant to a vaccine (4).

The question of a need for a third dose remains open. Some countries had decided to apply a booster for severely immunocompromised individuals, but it is still unknown if it would be necessary to the public in general. Therefore, this study aimed to measure SARS-CoV-2 spike 1-2 IgG antibodies in healthcare workers vaccinated with the complete two-dose regimen of BNT126b2 who received a third booster dose after six months of the second dose (12).

## Materials and methods

It was a prospective observational study that followed the STROBE guidelines (13). This study analyzed a subgroup of patients who completed the two-dose regimen of BNT126b2 at the beginning of 2021 in Monterrey, Nuevo Leon, Mexico. The study was approved by the local Institutional Review Board (Ref.:26022021-CN-1e-CI) and conducted per the Code of Ethics of the World Medical Association (Declaration of Helsinki) for experiments that involve humans.

The inclusion criteria for the patients’ recruitment were individuals of both genders between the age of 18 and 100 years old, who had accepted and signed an informed consent form and pretended to complete the BNT126b2 regimen. The patients were excluded if they did not complete the established vaccine regimen or received another SARS-CoV-2 vaccine.

The individuals were introduced to the research process, which consisted of a follow-up during an entire year through SARS-CoV-2 specific IgG antibodies measurement samples followed by questionnaires. After they agreed, they signed up for a written informed consent form. As this subgroup were healthcare employees, they were immunized against SARS-CoV-2 in the country with BNT126b2 at the beginning of 2021. Since this protocol was approved after the application of the first BNT, twenty-one days after receiving the first dose, the research team reached every participant to take a plasma sample for the IgG antibodies measurement and apply the first questionnaire. This questionnaire aimed to obtain the patient’s medical history, SARS-CoV-2 infections before and after being immunized and identify adverse events after the vaccine application.

After 21-28 days of receiving the second dose, the participants were scheduled to take the second IgG antibodies sample with its corresponding questionnaire similar to the one answered before. The third, fourth, and fifth IgG antibodies samples were planned to be taken three, six, and twelve months after completing the two-dose regimen of BNT126b2, respectively. The questionnaires applied in these samples aimed to know any suspicious or confirmed SARS-CoV-2 infections.

The plasma sampling required 10 to 15 ml of blood per venipuncture. These samples were kept in sample tubes with ethylenediaminetetraacetic acid (EDTA) as an anticoagulant, and they were stored at −80 °C. The equipment used by the laboratory personnel to analyze the samples was the LIAISON SARS-CoV-2 S1 / S2 IgG antibody detection kit (Italy). To determine the amount of specific anti-S1 and anti-S2 IgG antibodies against SARS-CoV-2 in plasma samples, the laboratory personnel used the chemiluminescence immunoassay (CLIA); which had a sensitivity of 97.4% (95% CI, 86.8-99.5) and a specificity of 98.5% (95% CI, 97.5-99.2). The results were reported as follows: <12.0 AU / ml was considered negative, 12.0 to 15.0 AU / ml was indeterminate, and > 15 AU / ml was positive.

In the three-month follow-up, there was a decrease in the amount of specific anti-S1 and anti-S2 IgG antibodies against SARS-CoV-2. This finding raised concern in the healthcare workers, leading them to seek a BNT126b2 boost shot out of the original BNT regimen by their own means. Furthermore, after the six months of the two-dose BNT162b2 regimen, a hundred and sixty-eight participants received a third booster shot. Given the situation, another blood sample was taken to the participants 21-28 days after the applied third boost.

The variables we analyzed were: age, gender, medical history, including confirmed SARS-CoV-2 diagnosis (confirmed either with a nasal swab or serologic tests before and after the BNT126b2 shots), and adverse events caused by any dose of BNT126b2. The analyzed biochemical variables were: SARS-CoV-2 quantitative antibodies from 21-28 days post-BNT126b2 first dose (S1), 21-28 days post-BNT126b2 second dose (S2), three-month follow-up after completing the two-dose BNT126b2 regimen (S3), and 1-7 days (S4) and 21-28 days post-BNT-boost (S5).

The researchers reviewed the quality control and the anonymization of the database. We evaluated descriptive statistics such as median, the interquartile range for quantitative variables, frequencies, and percentages for categorical variables. The group comparison was analyzed through Cochran’s Q test. The statistical program used was SPSS, version 2. The analysis was two-tailed. A p-value < 0.05 was considered statistically significant.

## Results

The recruited participants with a three-dose BNT126b2 regimen were 168, of which 113 (67.3%) were women. The mean (SD) age was 41 (10) years old. The most common comorbidity was obesity (44, 26.2%), followed by dyslipidemia (15, 8.9%) and hypertension (14, 8.3%), respectively. Table 1. shows the medical history of the participants stated in the first questionnaire. The time elapsed (SD) between the first and second BNT126b2 shots was 31 (4) days, and between the second and third dose was 166 (12) days.

**Table 1.** Medical history

| **Medical history (n=168)** | **Frequency (%)** |
| --- | --- |
| Obesity | 44 (26.2) |
| Dyslipidemia | 15 (8.9) |
| Hypertension | 14 (8.3) |
| Diabetes | 10 (6) |
| Prediabetes | 6 (3.6) |
| Smoking | 12 (7.1) |
| Pregnancy | 1 (0.6) |

Data are presented in frequencies and percentages.

According to the previous history of SARS-CoV-2 diagnosis, 95 (56.5%) of the participants reported never having the infection. The suspected cases of SARS-CoV-2 before vaccination were 4 (2.4%). Seventy-three (43.5%) participants stated a SARS-CoV-2 diagnosis any time before being vaccinated to 21-28 days after the applied third BNT shot, of which 52 (31%) reported it once, 17 (10.1%) twice, and 4 (2.4%) thrice. Before vaccination, healthcare workers with confirmed SARS-CoV-2 infection were 65 (38.7%) once and 7 (4.2%) twice. Participants with SARS-CoV-2 diagnosis between the first and second BNT shots were 3 (1.8%); between the second and third BNT shots, 1 (0.6%); and after the third dose, 0. Table 2 shows the history of SARS-CoV-2 infection in the participants in frequencies and percentages.

**Table 2.** History of SARS-CoV-2 infection

| **History of SARS-CoV-2 (n=168)** | **Frequency (%)** |
| --- | --- |
| Negative | 95 (56.5) |
| Suspected cases before vaccination | 4 (2.4%) |
| **Positive history any time between before vaccination to 21-28 days after the third dose** |  |
| Positive | 73 (43.5) |
| Once | 52 (31) |
| Twice | 17 (10.1) |
| Thrice | 4 (2.4) |
| **Positive history before vaccination** |  |
| Once before vaccination | 65 (38.7) |
| Twice before vaccination | 7 (4.2%) |
| **Positive history after initiating vaccination regimen** |  |
| Between first and second BNT shots | 3 (1.8%) |
| Between second and third BNT shots | 1 (0.6%) |
| After the third BNT shot | 0 (0) |

Data are presented in frequencies and percentages. The table is divided in positive history any time during the investigation, before vaccination, and after initiating the vaccination regimen.

The median (IQR) quantitative SARS-CoV-2 spike 1-2 IgG antibody titers obtained in the healthcare workers with no history of SARS-CoV-2 (n=95) was 2960 (2010) after the BNT162b2 third dose. In the participants with history of SARS-CoV-2 infection (n=73) the results were 1130 (4756) in S1; 2390 (2540) in S2; 377.5 (1144.75) in S3; 590 (1262) in S4; and 3090 (2080) in S5. Three months after the complete BNT162b2 scheme was noticed, the antibodies were reduced by 84.8% and 84.4% in negative and positive SARS-CoV-2 infection participants, respectively. Table 3. presents the median (IQR) of the quantitative SARS-CoV-2 IgG antibody titers of the participants divided into two groups, depending on their history of SARS-CoV-2 infection.

**Table 3.** Quantitative SARS-CoV-2 spike 1-2 IgG antibody titers against SARS-COV-2 in the positive and negative history SARS-CoV-2 infection participants

| **History of SARS-CoV-2 (n=168)** | **21-28 days after first BNT shot (S1) (QIR)** | **21-28 days after second BNT shot (S2) (QIR)** | **Three months after the second BNT shot (S3) (QIR)** | **1-7 days after the third BNT boost (S4) (QIR)** | **21-28 days after the third BNT boost (S5) (QIR)** |
| --- | --- | --- | --- | --- | --- |
| **Negative (n=95)** | 103 (77.6) | 1350 (1224) | 205 (149) | 379 (645.5) | 2960 (2010) |
| **Positive (n=73)** | 1130 (4756) | 2390 (2540) | 377.5 (1144.75) | 590 (1262) | 3090 (2080) |
| **p-value** | <0.001 | <0.002 | 0.001 | 0.011 | 0.377 |

Data are presented in median and interquartile ranges. The participants were divided into two groups, those with a negative and positive history of SARS-CoV-2.

Figure 1 shows box plot schemes of the quantitative SARS-CoV-2 spike 1-2 IgG antibody titers against SARS-COV-2 to compare the results between the groups with negative or positive SARS-CoV-2 history.

**Figure 1.**
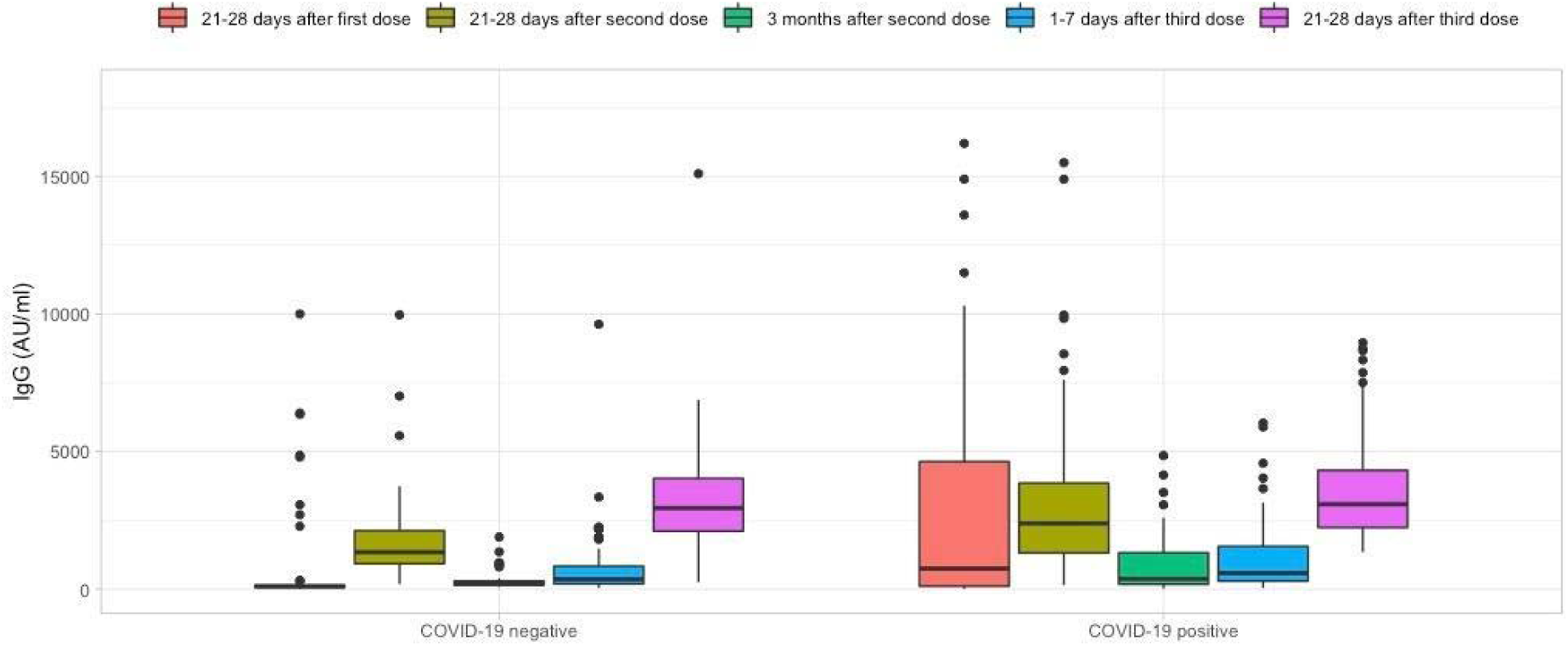
Box plot scheme of the quantitative SARS-CoV-2 spike 1-2 IgG antibody titers by SARS-CoV-2 history. Quantitative SARS-CoV-2 spike 1-2 IgG titers after the first and second dose, three months, 1-7 days after the third dose, and 21-28 days following a booster dose of BNT162b2 vaccine, divided into two groups according to the SARS-CoV-2 history.

Side effects were reported after the first, second, and third BNT shots; the less total number of side effects were reported after the third BNT162b2 dose (n=140, 83%; n=133, 79.1%; and n=97, 57.7%, respectively) (p<0.001). The first and second BNT dosages’ side effects appeared most frequently in the first four hours after application (n=116, 82.8%; and n=59, 44.4%, respectively), while after the third shot were reported 5 to 24 hours after (n=56, 57.7%). The severity was considered very mild after the first and second BNT shots (n=95, 67.8%; and n=57, 42.8%, respectively), and mild after the third BNT boost (n=35, 36%).

The most common side effect after the three applied shots was pain at the injection site (n=131, 93.6%; n=119, 89.5%; and n=82, 84%, respectively), followed by headache (n=51, 36.4%; n=58, 43.6%; and 43, 44.3%, respectively), and tiredness (n=38, 27.1%; n=54, 32.1%; and n=45, 46.4%, respectively). When comparing the side effects in each dose, tiredness, myalgias, arthralgias, fever, and adenopathy were proportionally higher following the third dose, compared to the two-dose regimen (p<0.05). Table 4 shows the side effects reported by the participants in frequencies and percentages, as well Table 5 shows the time of appearance and severity.

**Table 4.** Side effects related to each BNT162b2 dose applied.

| Side effects<br>(n=168) | First dose (%) | Second dose<br>(%) | Third dose (%) | p-value |
| --- | --- | --- | --- | --- |
| **Present** | 140 (83.3) | 133 (79.1) | 97 (57.7) | < 0.001 |
| **Symptoms** |  |  |  |  |
| **Pain at the**<br>**injection site** | 131 (93.6) | 119 (89.5) | 82 (84.5) | 0.204 |
| **Headache** | 51 (36.4) | 58 (43.6) | 43 (44.3) | 0.459 |
| **Tiredness** | 38 (27.1) | 54 (40.6) | 45 (46.4) | 0.03 |
| **Myalgia** | 12 (8.6) | 38 (22.6) | 27 (27.8) | 0.001 |
| **Local edema or**<br>**erythema** | 11 (6.5) | 20 (15) | 15 (15.5) | 0.421 |
| **Arthralgia** | 8 (5.7) | 37 (22) | 26 (26.8) | 0.001 |
| **Fever** | 4 (2.9) | 4 (3) | 11 (11.3) | 0.003 |
| **Insomnia** | 5 (3) | 3 (2.3) | 5 (5.2) | 0.561 |
| **Palpitations** | 4 (2.4) | 7 (5.3) | 7 (7.2) | 0.641 |
| **Thoracic pain or oppression** | 5 (3) | 6 (4.6) | 5 (5.2) | 0.846 |
| **Nasal congestion** | 4 (2.9) | 17 (12.8) | 8 (8.2) | 0.047 |
| **Adenopathy** | 1 (0.7) | 3 (2.3) | 14 (14.4) | 0.001 |
| **Hypotension** | 1 (0.7) | 2 (1.5) | 2 (2.1) | 0.368 |
| **Pruritus** | 1 (0.7) | 4 (3) | 3 (3.1) | 0.247 |
| **Ocular pain** | 2 (1.7) | 0 | 0 | 0.368 |
| **Nausea** | 4 (2.9) | 11 (8.3) | 6 (6.2) | 0.035 |
| **Diarrhea** | 0 | 5 (3.8) | 4 (4.1) | 0.135 |

Data are presented in frequencies and percentages, showing the presence of side effects, as well as the most common symptoms, time of appearance, and severity. Cochran's Q test was performed for comparison between the side effects of each dose. A p-value $\leq 0.05$ was considered statistically significant.

**Table 5.**
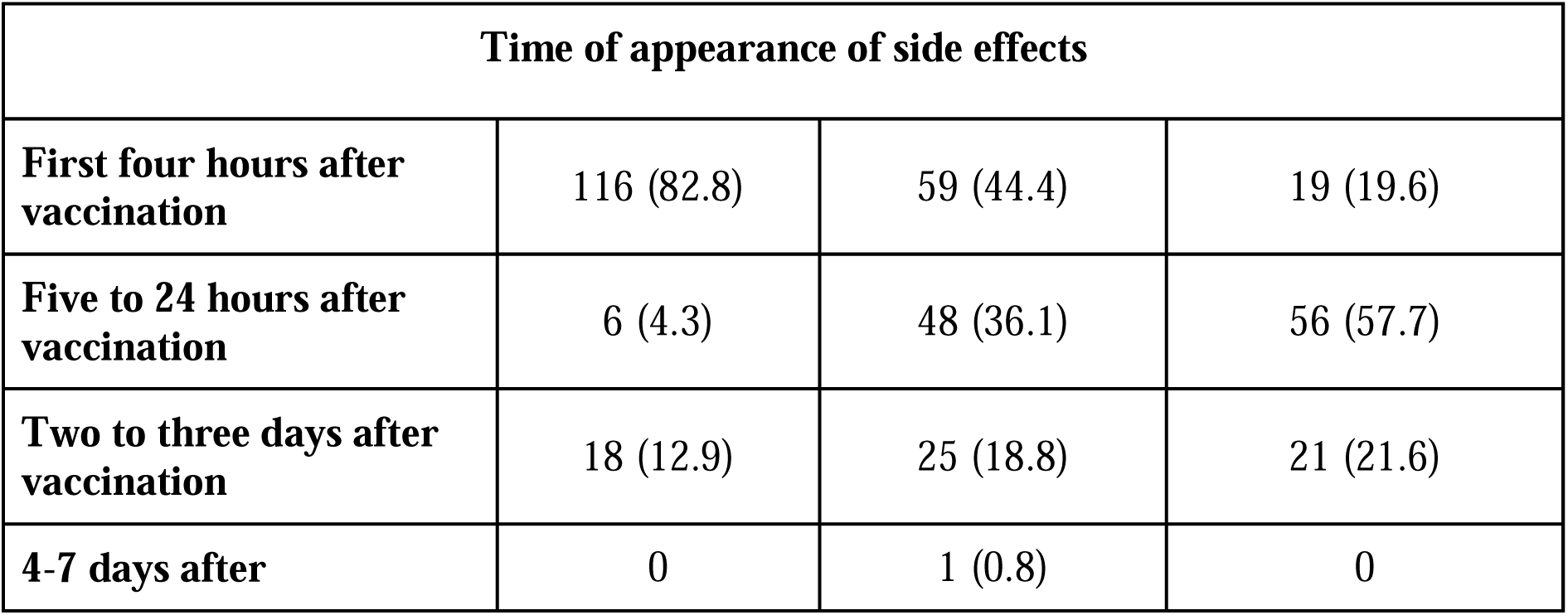
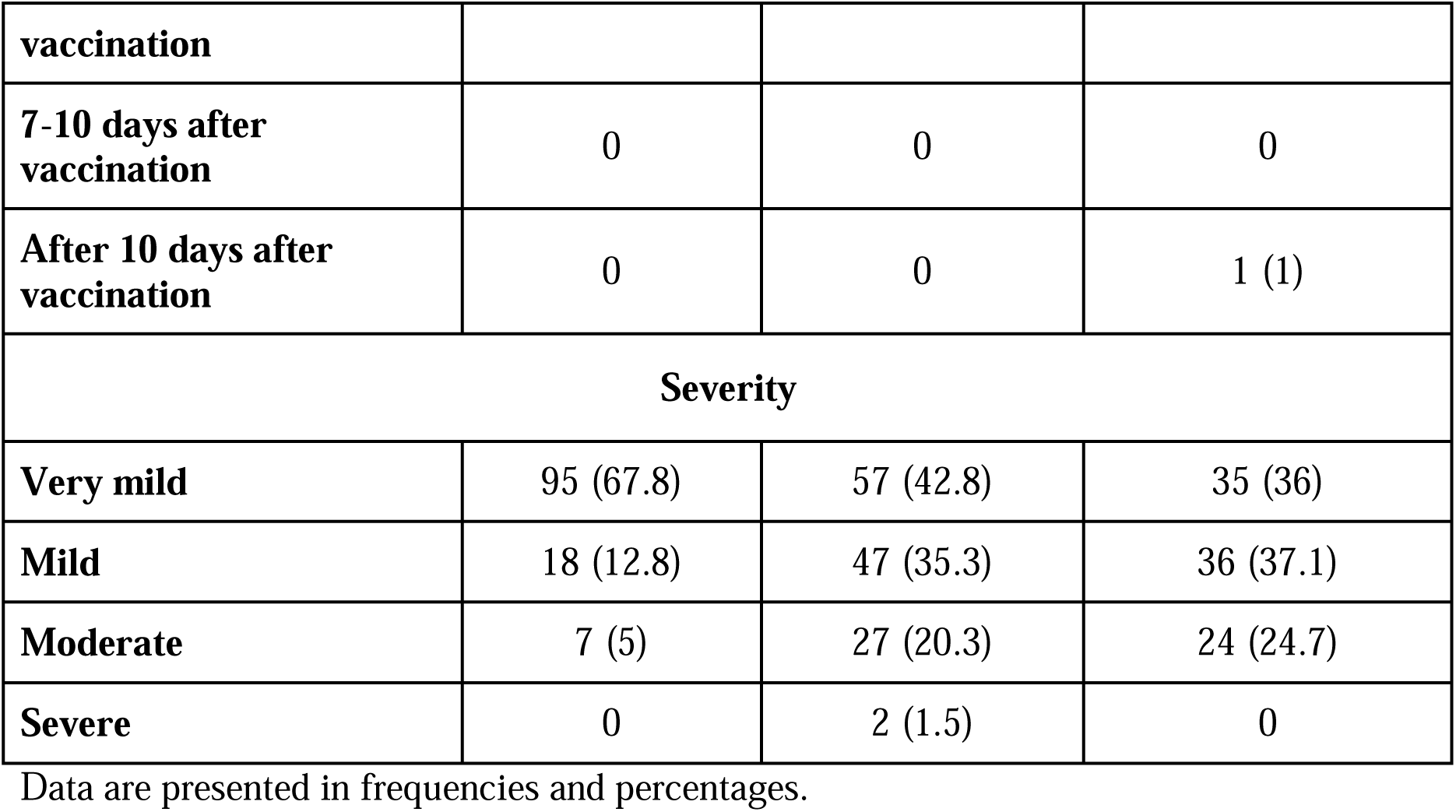
Time of appearance and severity of side effects related to the BNT162b2 doses

| **Time of appearance of side effects** |  |  |  |
| --- | --- | --- | --- |
| **First four hours after vaccination** | 116 (82.8) | 59 (44.4) | 19 (19.6) |
| **Five to 24 hours after vaccination** | 6 (4.3) | 48 (36.1) | 56 (57.7) |
| **Two to three days after vaccination** | 18 (12.9) | 25 (18.8) | 21 (21.6) |
| **4-7 days after** | 0 | 1 (0.8) | 0 |
| **vaccination** |  |  |  |
| **7-10 days after vaccination** | 0 | 0 | 0 |
| **After 10 days after vaccination** | 0 | 0 | 1 (1) |
| **Severity** |  |  |  |
| **Very mild** | 95 (67.8) | 57 (42.8) | 35 (36) |
| **Mild** | 18 (12.8) | 47 (35.3) | 36 (37.1) |
| **Moderate** | 7 (5) | 27 (20.3) | 24 (24.7) |
| **Severe** | 0 | 2 (1.5) | 0 |

Data are presented in frequencies and percentages.

## Discussion

The purpose of this study was to analyze the quantitative SARS-CoV-2 spike 1-2 IgG antibody titers throughout six months after completing the established BNT126b2 vaccination regimen and 21-28 days following a third BNT126b2 dose applied six months after the second dose. After three months of the second dose’s application, the antibodies decreased compared to the titers seen 21-28 days after the second dose. In addition, the sample taken 1-7 days after the third dose, equivalent to the sample after six months of the complete regimen, the antibody titers were lower than the ones reported following 21-28 days of the second dose.

Previous studies have demonstrated a decline in the quantitative SARS-CoV-2 spike 1-2 IgG antibody titers after completing the BNT126b2 scheme in non-immunocompromised patients, regardless of their previous SARS-CoV-2 history. Alejo Erice et al. reported a reduction of 58% of the quantitative SARS-CoV-2 spike 1-2 IgG antibody titers after three months (14).

In addition, the study conducted by Julien Favresse et al. informed a decrease in the antibody titers between 56 to 90 days after the second BNT162b2 vaccine being observed in both seronegative and seropositive participants at the beginning of the study (15). Our study demonstrated to be consistent with these previous investigations because of the decrease of quantitative SARS-CoV-2 spike 1-2 IgG antibody titers observed in the third sample. Three months after the complete BNT162b2 scheme, the antibodies were reduced by 84.8% and 84.4% in negative and positive SARS-CoV-2 infection participants, respectively.

At 21-28 days following the administration of the third BNT162b2 boost, the quantitative SARS-CoV-2 spike 1-2 IgG antibody titers increased drastically. The non-immunocompromised participants with a negative SARS-CoV-2 infection increased the antibody titers more than six times the previous result obtained in the sample taken 1-7 days after the third dose, while positive SARS-CoV-2 infection, more than four times.

Therefore, in a study where heart transplant recipients were vaccinated with a third BNT162b2 dose, Yael Peled, et al. informed a 3-fold increase in the anti-receptor binding domain antibodies, despite immunosuppressive therapy (16). Additionally, according to Yinon M Bar-On et al., following a third BNT162b2 booster dose administered in an Israeli group of people older than 60 years old, the rates of confirmed SARS-CoV-2 infection and severe illness were diminished compared to a control group with the original BNT162b2 two-dose regimen (17). Given these situations associated with our findings, a three homologous BNT162b2 dose regimen applied six months after the second shot provides a beneficial effect regarding a positive humoral immune response against the SARS-CoV-2 infection.

During the follow-up after the six months prior to the third dose, we noticed higher results of quantitative SARS-CoV-2 spike 1-2 IgG antibody titers in the participants with a positive history of SARS-CoV-2, in comparison to the ones with a negative history. Interestingly, in the results obtained 21-28 days after the third booster dose, the difference between the participants with positive or negative SARS-CoV-2 history was not perceptible as with the previous two shots. This means that after the third dose, participants with a negative SARS-CoV-2 history have the same level of humoral immunity as the ones exposed previously to a SARS-CoV-2 infection.

The main concern about the third dose’s application was the lack of knowledge of the possible adverse effects, which this additional BNT162b2 dose could cause. Yael Peled et al. reported that 67% of the participants with a third BNT162b2 boost reported at least one side effect of mild severity. The most common side effects were pain at the injection site, tiredness, and headache, with no hospitalizations or emergency room (ER) visits (16). Our study also found that the side effects associated with the third dose were mild, and the most frequent were pain on the injection site, tiredness, and headache, with no severe complications such as hospitalization or ER visits. In comparison to the other shots, the presence of side effects after the third dose was the least reported, given by a decrease of 30.7% and 27.0% compared to those reported in the first and second BNT162b2 shots, respectively.

To the best of our knowledge, no previous studies show the response of quantitative SARS-CoV-2 spike 1-2 IgG antibody titers to a third BNT162b2 boost in non-immunocompromised healthy patients. We believe this study may contribute to the knowledge of the healthcare community because we observed a positive humoral immune response to the third BNT162b2 dose in our non-immunocompromised healthcare workers. This can lead to better protection for the healthcare workers and the groups vulnerable to severe illness by SARS-CoV-2.

Furthermore, the approval of the third BNT162b2 dose and its proven efficacy could lead to better control of the SARS-CoV-2 pandemic, providing vulnerable groups greater immunogenicity against the virus with this additional dose. Consequently, the number of confirmed cases in Mexico would decrease as well as hospitalizations by this agent.

A limitation to our study was that the recruited group did not have a baseline sample taken before the first BNT162b2 dose because our protocol was approved after the health workers had received the first dose. However, we believe that this study is significant because of the non-immunocompromised population that we used. Another limitation was the analysis of the third dose’s side effects only in the short-term (21-28 days after the booster dose). We will continue following this cohort group to see any long-term side effects related to the vaccine, but we just reported the short-term side effects for this study.

## Conclusion

The third BNT162b2 booster dose has proven to increase the quantitative SARS-CoV-2 spike 1-2 IgG antibody titers with no severe side effects in the short-term. Despite its positive response, we must continue studying this cohort to see if the antibody titers are maintained, and the SARS-CoV-2 infections decrease, as well as a severe illness caused by the same agent.

BNT162b2 Pfizer/BioNTech is one of the most studied vaccines due to its efficacy and worldwide application. Due to the approval of its third dose by the CDC and FDA, further investigations of BNT162b2 regarding this new vaccination scheme will provide critical knowledge to the medical field that will contribute to better control of the pandemic. Our cohort study might be considered significant because no other studies that measured quantitative SARS-CoV-2 spike 1-2 IgG antibody titers in non-immunocompromised healthcare workers were found when this article was written.

We will continue pursuing our study for further findings in order to provide more relevant information to contribute to the control of the SARS-CoV-2 pandemic.

## Data Availability

All data produced in the present study are available upon reasonable request to the authors

## Author Approval

all authors read an approved the final version of the manuscript

## Competing Interests

The authors have declared no competing interest

## Funding

The research was supported by private funding provided by the hospital. No external funding was used.

## Ethics statement

Ethics committee/local Institutional Review Board from the school of Medicine from Universidad de Monterrey gave ethical approval: Ref.:26022021-CN-1e-CI

## Acknowledgments

We thank the laboratory team, human resources, call center, technology, and maintenance personnel of Hospital Clinica Nova, for helping in the logistics of this research.

## Notes

### Clinical Trial

NA

